# Returning Individual-Level Urgent or Emergent Research Results to Participants: The Project Baseline Health Study Experience

**DOI:** 10.1101/2024.09.19.24313995

**Authors:** Neha Pagidipati, Brooke Heidenfelder, Lydia Coulter Kwee, Fatima Rodriguez, Ranee Chatterjee, Kishan S. Parikh, Michel G. Khouri, Jennifer Stiller, Julie Eckstrand, P. Kelly Marcom, Priyatham S. Mettu, Glenn J. Jaffe, Sumana Shashidhar, Susan Swope, Susan Spielman, Elizabeth Fraulo, L. Kristin Newby, Pamela Douglas, Charlene Wong, Robert Green, Scarlet Shore, Jessica Mega, Adrian Hernandez, Paul Campbell, Kenneth W. Mahaffey, Svati H. Shah, Project Baseline Health Study Group

**Affiliations:** Division of Cardiology, Department of Medicine, Duke University School of Medicine, Durham, NC, USA; Duke Clinical and Translational Science Institute, Duke University School of Medicine, Durham, NC, USA; Duke Molecular Physiology Institute, Duke University School of Medicine, Durham, NC, USA; Division of Cardiovascular Medicine, Department of Medicine, and the Cardiovascular Institute, Stanford University School of Medicine, Stanford, CA, USA; Division of General Internal Medicine, Department of Medicine, Duke University School of Medicine, Durham, NC, USA; WakeMed, Raleigh, NC, USA (present affiliation); Duke Clinical and Translational Science Institute, Duke University School of Medicine, Kannapolis, NC, USA; Duke Cancer Institute, Duke University School of Medicine, Durham, NC, USA; Department of Ophthalmology, Duke University School of Medicine, Durham, NC, USA; Stanford Center for Clinical Research, Department of Medicine, Stanford University School of Medicine, Stanford, CA, USA; Duke Clinical Research Institute, Duke University School of Medicine, Durham, NC, USA; Department of Pediatrics, Duke University School of Medicine, Durham, NC, USA; Mass General Brigham, Broad Institute, Ariadne Labs and Harvard Medical School, Boston, MA, USA; Verily Life Sciences, Inc., South San Francisco, CA, USA; Sanger Heart & Vascular Institute Concord, Concord, NC, USA; Stanford Cardiovascular Institute, Stanford University School of Medicine, Stanford, CA, USA

**Keywords:** return of results, incidental findings, prospective studies, cardiovascular disease, lung disease, diabetes

## Abstract

**Background:** Returning results to research participants is increasingly recognized as an ethical mandate, yet little is known about best practices to optimally communicate urgent or emergent results.

**Methods:** The Project Baseline Health Study (PBHS) was a prospective observational cohort study of 2,502 participants enrolled from 2017-2019 and followed through 2023. The cohort represented a broad spectrum of health and disease, enriched to include participants with an elevated risk for breast or ovarian cancer, lung cancer, and/or atherosclerotic cardiovascular disease (60% of participants). Urgent or emergent results were returned during or after the baseline visit from vital signs; clinical laboratory testing; and ocular, cardiovascular, and pulmonary imaging.

**Results:** Among 2,002 participants in this analysis, 39.7% had at least one urgent or emergent finding returned, representing a total of 1,159 results returned over 3 years. The most commonly returned results were eye findings (n=246), pulmonary nodules (n=159), abnormal stress echocardiograms (n=123), abnormal rest electrocardiograms (bradycardia) (n=74), and lung parenchyma findings (n=55). Participants with urgent or emergent incidental findings were older (mean [SD] 58.0 [16.2] years vs. 48.0 [16.6] years) with a greater burden of cardiovascular, metabolic, or cancer comorbidities than those without urgent or emergent incidental findings.

**Conclusions:** This report from the PBHS study is one of the first to describe a process to systematically return urgent or emergent results to research participants. This process led to the successful return of clinically important results to participants but also required significant time and effort from study clinicians and staff.

**ClinicalTrials.gov Identifier:** NCT03154346

## INTRODUCTION

The return of individual results to research participants is a key component of the overarching shift in research toward the democratization of data and open science (1). The arguments favoring the return of individual-level results are substantive and rooted in an ethical case for respecting all persons and their self-determination and not treating participants as simply a means to an end to generate scientific data (2). Indeed, participants have a right to their own data and should be considered partners in research (1,3). Beyond the ethical argument for return of results is the empirical argument that participants have a clear desire to have their own data returned to them. A survey of over 2,500 participants in various types of research studies across the United States (U.S.) revealed that most thought that receiving results would be valuable (78.5%), should be expected (71.7%), and would make them more likely to participate in research (72.4%) and trust researchers (70.3%) in the future (4).

Despite the many arguments in favor of returning research results to participants, little is known about optimal approaches for returning results that are considered clinically urgent or emergent (1). A consensus study report from the National Academies of Science, Engineering, and Medicine provided 12 recommendations to return results to research participants (5). However, none of these recommendations provide specific guidance on how to structure a process to optimally return urgent or emergent findings.

Here, we describe a process developed for the Project Baseline Health Study (PBHS) to return urgent or emergent findings to research participants. The PBHS study enrolled a diverse population across age, sex, race, and comorbidities to conduct deep longitudinal clinical and molecular phenotyping over 4 years at multiple sites in the U.S. (6). Assessments included eye imaging, echocardiogram and stress echocardiogram, chest computed tomography (CT) for coronary calcium, clinical laboratory testing, and molecular profiling. A Return of Results (RoR) Committee was established to develop a standardized approach to return results to participants, including urgent or emergent findings. In this analysis, we assessed the proportion and characteristics of PBHS participants who had incidental findings returned. In addition to reporting these results, we describe the RoR Committee protocols, and we present a model designed to predict which participants were most likely to have urgent or emergent results to understand how to better plan for resource allocation. While specific to the PBHS study, this model outlines an approach that can be replicated in other studies to help with resource allocation.

## METHODS

### PBHS Overall Design and Study Participants

The design of the PBHS study has been described previously (6). In brief, the PBHS study was designed to establish a contemporary reference health state and to study transitions into various states of disease through in-depth, longitudinal, multi-dimensional phenotyping. Individuals selected to participate were chosen to create a broad cohort, spanning the health spectrum and reflecting the demographic and clinical characteristics of U.S. Census data (6). In addition to this broad enrollment, the cohort was enriched to include participants with an elevated risk for breast or ovarian cancer, lung cancer, and/or atherosclerotic cardiovascular disease (60% of participants) (6). In total, 2,502 participants were enrolled across four sites in the U.S., including Duke University in Durham, North Carolina; Duke University’s research site in Kannapolis, North Carolina; Stanford University; and the California Health and Longevity Institute (CHLI). Participants were followed annually with in-person visits through 2023. For this analysis, only data from the baseline visits at the Duke-Durham, Duke-Kannapolis, and Stanford sites were included (N=2,002). The results from CHLI participants were not included as they did not have on-site clinical reads performed for several assessments; therefore, the timing and process for returning results was different from the other sites.

Participants were consented for participation and underwent an extensive 2-day set of baseline assessments that included vital signs, imaging (ophthalmologic, chest X-ray, echocardiography, electrocardiography, stress echocardiography, and coronary calcium CT scanning with inclusion of lung fields), biospecimen collection, medical history, surveys, arterial brachial index measurements, and pulmonary function tests. The study was approved by two institutional review boards. Participants provided written informed consent before participation, and the study observed the privacy rights of all participants.

### Return of Results Protocols

The RoR Committee protocols focused on clinical results performed in Clinical Laboratory Improvement Amendments (CLIA)-certified laboratories and/or testing with clear clinical implications performed in a clinical environment. The RoR Committee considered the implications of returning research-grade assessments and determined that molecular profiling (e.g. DNA sequencing, proteomics, gene expression, microbiome) was considered research and since it was not conducted in a CLIA lab, these results would not be returned. Participants did not have the option to decline receiving study results.

The assessments performed at baseline for the study participants are outlined in **Table 1**. The results from the following testing modalities performed at enrollment were eligible to be returned to participants: vital signs; clinical laboratory testing (**Supplementary Table 1**); chest CT scan; chest X-ray; resting electrocardiogram; resting and stress echocardiogram; and ophthalmologic findings from keratometry/corneal topography, optical coherence tomography, and retinal photography. A digital health device was worn for at least 10 hours each day, and average individual-level results from this device (e.g. daily step count) were returned to participants after enrollment in the PBHS study was completed; however, because these results were not considered urgent or emergent, they were not included in this analysis.

**Table 1.** Baseline Assessments Performed at Enrollment.

| **Clinical** | **Biospecimen** | **Imaging** | **Self-Report** |
| --- | --- | --- | --- |
| Ankle-brachial index* | Blood—clinical laboratory testing* | Chest X-ray | Patient Health Questionnaire-9 (PHQ-9)* |
| Medical history | Saliva | Coronary calcium scan (chest CT) | Generalized Anxiety Disorder-7 (GAD-7)* |
| Review of systems | Stool | Resting echocardiogram | EuroQol 5 Dimension 5 Level (EQ-5D-5L) |
| Physical performance | Swabs (buccal, retroauricular, nares, oral) | Stress echocardiogram | Behavioral Risk Factors Surveillance System (BRFSS)—Adverse Childhood Experience Module (ACE) |
| Pulmonary function testing* | Urine | Resting electrocardiogram | Sheehan Disability Scale (SDS) |
| Visual performance |  | Keratometry/corneal topography | World Health Organization Disability Assessment Schedule 2.0 12-item version (WHODAS 2.0) |
| Vital signs* |  | Optical coherence tomography |  |
|  |  | Retinal photography |  |

\*Eligible for return of results
CT, computed tomography.

The RoR Committee worked with PBHS site and core lab clinicians with relevant expertise, including non-invasive imaging cardiologists, ophthalmologists, and pathologists, to develop a color flag system (**Figure 1**) for incidental findings on each assessment modality to determine which results should be returned and the level of timeliness after assessment. For each modality, potential findings were enumerated and categorized according to perceived clinical urgency. Findings considered critical with time-sensitive clinical actionability (i.e. emergent) were assigned a red flag, whereas those considered important and clinically actionable but not as time-sensitive (i.e. urgent) were assigned an orange flag. Results outside of the expected range and clinically actionable but not urgent, or of uncertain clinical or analytic validity, were assigned a yellow flag. Finally, those results within the expected range were assigned a green flag.

**Figure 1.**
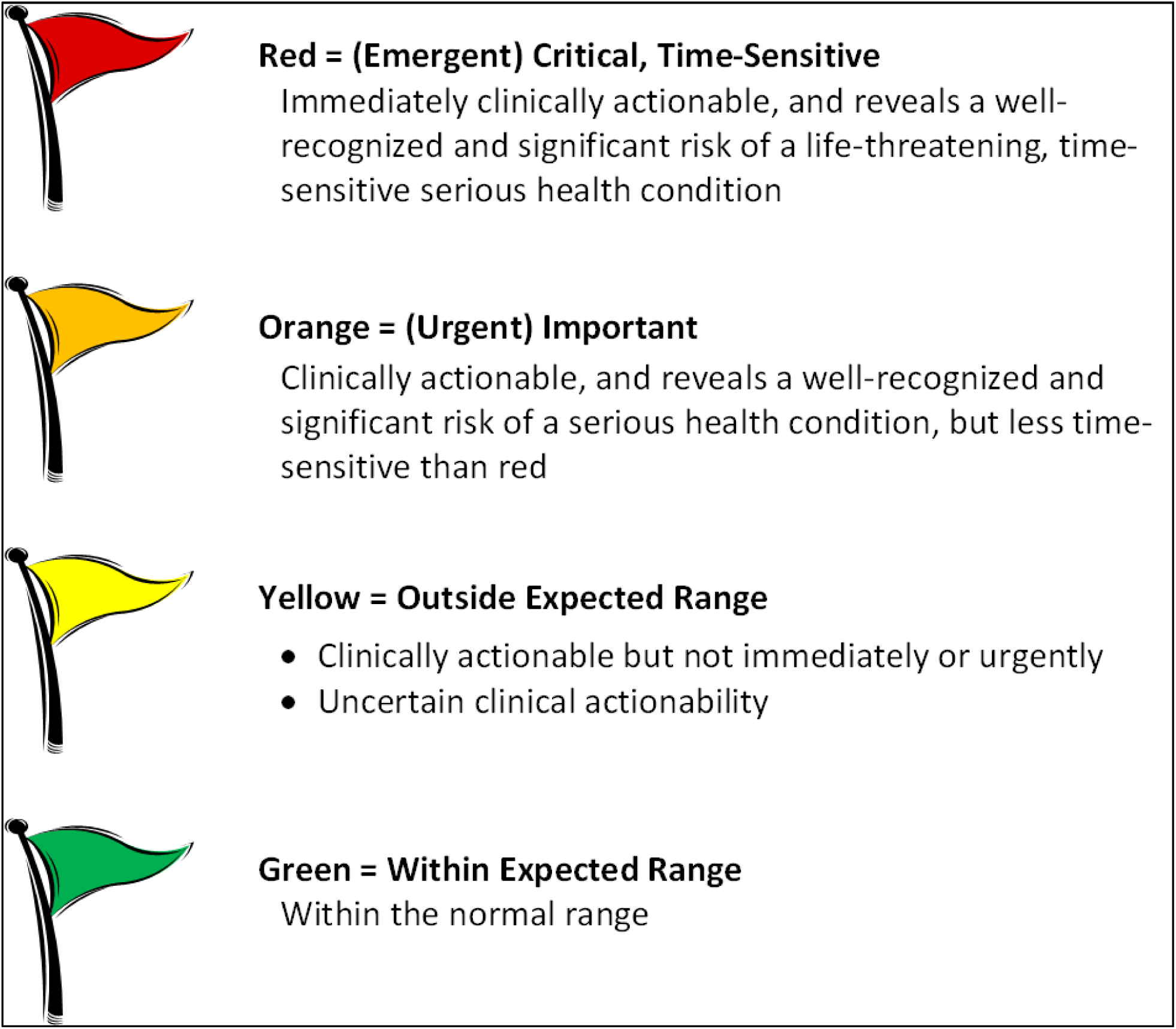
Return of Results Flag System.

Some results were available during the 2-day on-site study visit while other results were observed after the study visit when the results became available. Vital signs and abnormal findings on resting or stress electrocardiogram or echocardiogram (recognized by the technicians performing the test and confirmed by the on-site clinician) were often available during the study visit, and the remainder of findings were observed afterward. This necessitated two workflows for returning results, as outlined in **Figure 2A** and **Figure 2B**.

**Figure 2.**
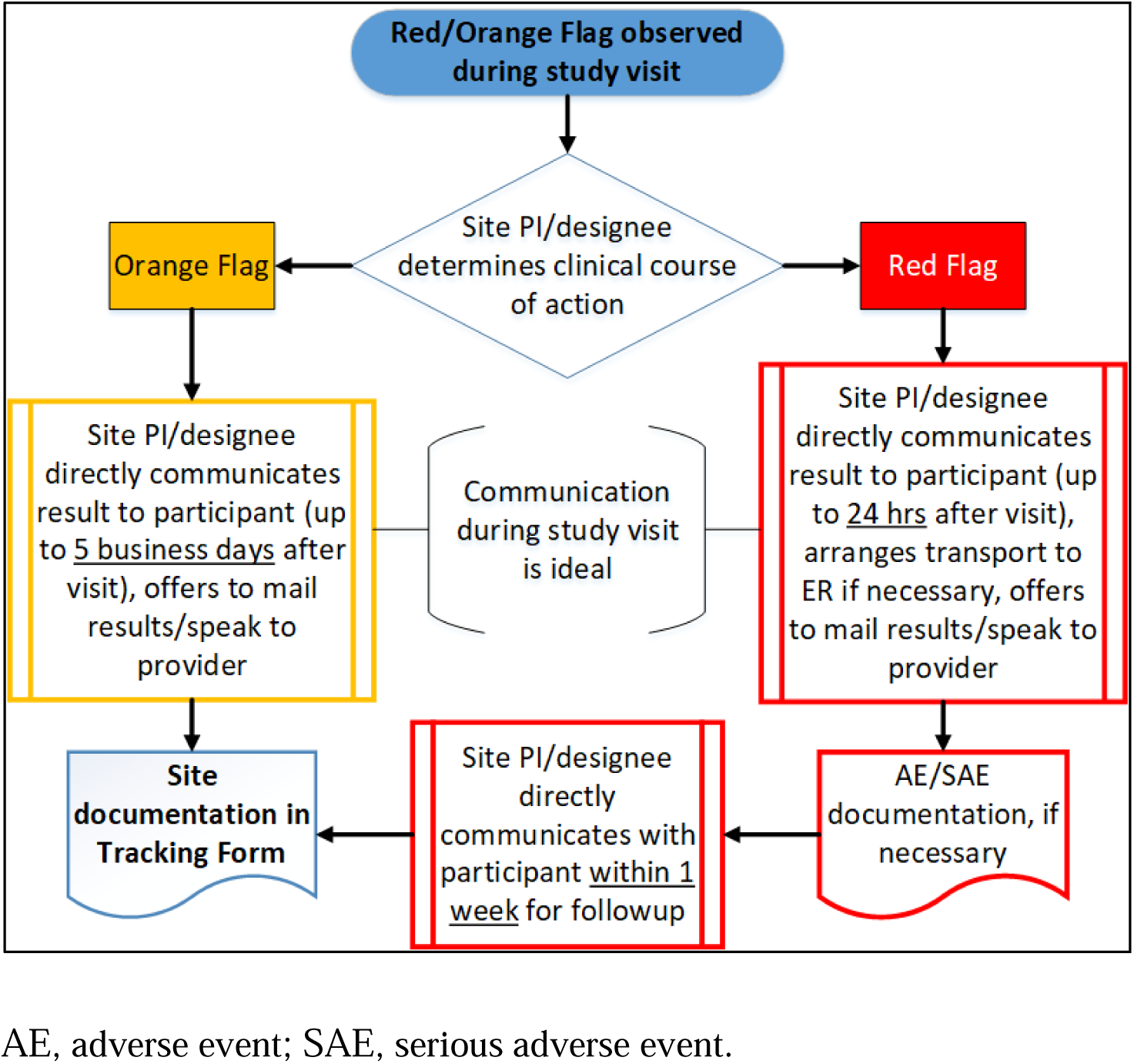
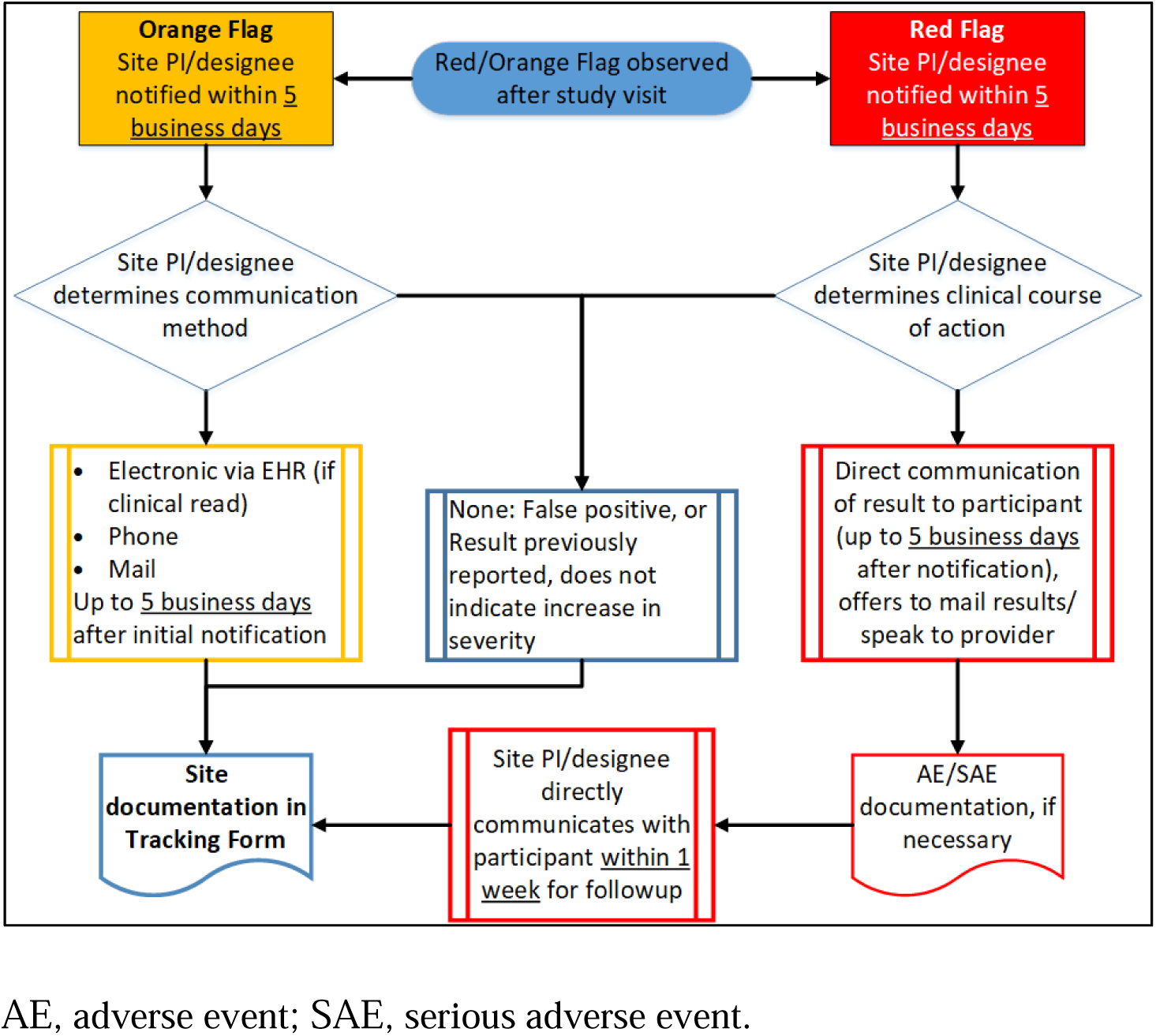
Return of Results Process. (A) Flag Observed During Study Visit. (B) Flag Observed After Study Visit.

Results were returned by site principal investigators (all practicing physicians) or their designees, all of whom were study physicians or advanced practice providers. Because of the volume of results requiring return, an “on call” system was set up in which rotating clinicians were assigned responsibility for returning urgent or emergent results in a given week.

### Return of Abnormal Results Identified During a Study Visit

If an emergent result was observed during the study visit, the responsible study clinician directly communicated this to the participant while they were still on-site, or within 24 hours of the abnormal result if that was not possible. Additionally, the study clinician arranged transport to the emergency department if necessary and offered to communicate with the participant’s clinician. The RoR Committee decided not to include return of results to participant clinicians unless requested by the participant, in part based on feedback from clinician groups. Within 7 days of the study visit, site designees contacted the participant for follow-up.

If an urgent result was observed during the study visit, a similar process was followed, except that the clinician was allowed 5 days to communicate the result to the participant, and there was no follow-up with the participant. For both emergent and urgent results, initial contact included asking the participant if the finding was already known to them and if they planned to seek follow-up care. As noted in the informed consent document, the study did not cover the costs of any health care required by the participant following the return of results.

### Return of Abnormal Results Identified Outside of a Study Visit

If an urgent or emergent result was identified after the study visit, site staff immediately informed the site physician. Initially, the RoR Committee allowed 24 hours for the return of emergent results identified after the visit and 5 days for the return of urgent results. However, this period was revised to 5 days for emergent results and 30 days for urgent results after the committee recognized the large burden of abnormal results and the clinical implications of these abnormal but not urgent abnormalities. For emergent findings, in addition to contacting the participant with the result, clinicians also offered to communicate with the participants’ primary clinician. Site staff re-contacted the participants at 1 week for follow-up. For urgent results, the site physician had the option to call, mail, or send the participant a message through the electronic health record (EHR) within 30 days of notification of the abnormal result.

When returning results to participants, site clinicians were instructed to relay the result itself and the necessity or urgency of follow-up. Site clinicians also asked if this finding was already known by the participant and whether they planned to seek follow-up care. However, because the site physicians did not have a role in the participant’s clinical care and were not aware of all the participant’s known comorbid conditions or symptoms, they were not permitted to give clinical advice (i.e. next steps for testing or management, implications for prognosis, etc.). Participants without an established clinician were given phone numbers for local primary care clinics and/or subspecialists as needed, including clinics that provided care to patients without health insurance.

### Statistical Methods

The number of urgent and emergent results are presented as simple counts along with the demographic and clinical characteristics of participants with abnormal results. Further, we developed a model that predicted which individuals were most likely to have an urgent or emergent result. The model included a set of broadly available clinical and laboratory variables (age, sex, race and ethnicity, smoking status, pulse, body mass index [BMI], blood pressure, lipids, estimated glomerular filtration rate [eGFR], HbA1c) and self-reported medical history (cancer, chronic kidney disease [CKD], hypertension, hyperlipidemia, stroke, coronary artery disease [CAD], chronic obstructive pulmonary disease [COPD], diabetes, heart failure, prior myocardial infarction [MI], and peripheral vascular disease). A LASSO regression model was built using the training set data, with the tuning parameter selected using 10-fold cross-validation with data randomly split into distinct training (60%) and test (40%) sets, each with similar proportions of urgent or emergent findings. We applied the final model to the test set and assessed the model’s accuracy, performance, and fit.

## RESULTS

From 2017-2019, a total of 2,002 participants underwent enrollment visits in the PBHS study at the Duke-Durham, Duke-Kannapolis, and Stanford sites (**Table 2**). The mean age of the participants was 52.0 years, and 55.5% of the participants were female. The racial and ethnic distribution of the participants was 62.6% White, 17.9% Black, 10.2% Asian, and 10.5% Hispanic. Overall, 3.9% had known CAD; 30.4% had a self-reported history of hypertension; 24.1% had hyperlipidemia; and 13.1% had type 2 diabetes mellitus. Reflecting the study design (6), 20.1% were at high risk for cardiovascular disease; 11.7% were at high risk for lung cancer; and 25.7% of female participants were at high risk for breast or ovarian cancer.

**Table 2.** Baseline Characteristics of PBHS Participants Overall and Stratified by the Presence or Absence of an Urgent or Emergent Incidental Finding.

|  | **Overall study cohort** | **Urgent or emergent finding** | **No urgent or emergent finding** |
| --- | --- | --- | --- |
| n | 2002 | 794 (39.7%) | 1208 (60.3%) |
| Site (%) |  |  |  |
| Duke-Durham | 485 (24.2) | 196 (24.7) | 289 (23.9) |
| Duke-Kannapolis | 515 (25.7) | 228 (28.7) | 287 (23.8) |
| Stanford | 1002 (50.0) | 370 (46.6) | 632 (52.3) |
| Age (mean [SD]) | 52.0 (17.2) | 58.0 (16.2) | 48.0 (16.6) |
| Sex (% Male) | 891 (44.5) | 382 (48.1) | 509 (42.1) |
| Self-reported race (%) |  |  |  |
| Black or African American | 358 (17.9) | 157 (19.8) | 201 (16.6) |
| White | 1254 (62.6) | 531 (66.9) | 723 (59.9) |
| Asian | 204 (10.2) | 54 (6.8) | 150 (12.4) |
| Native Hawaiian or Other Pacific Islander | 17 (0.8) | 9 (1.1) | 8 (0.7) |
| American Indian or Alaska Native | 23 (1.1) | 6 (0.8) | 17 (1.4) |
| Other Race | 146 (7.3) | 37 (4.7) | 109 (9.0) |
| Hispanic, Latino, or Spanish origin (%) | 210 (10.5) | 58 (7.3) | 152 (12.6) |
| BMI (mean [SD]) | 28.9 (7.0) | 29.5 (7.2) | 28.5 (6.8) |
| Smoking status (%) |  |  |  |
| Current | 298 (14.9) | 140 (17.6) | 158 (13.1) |
| Former | 466 (23.3) | 220 (27.7) | 246 (20.4) |
| Never | 1238 (61.8) | 434 (54.7) | 804 (66.6) |
| Hypertension (%) | 609 (30.4) | 319 (40.2) | 290 (24.0) |
| Hyperlipidemia (%) | 483 (24.1) | 260 (32.7) | 223 (18.5) |
| Type 2 diabetes (%) | 263 (13.1) | 159 (20.0) | 104 (8.6) |
| Coronary artery disease (%) | 79 (3.9) | 52 (6.5) | 27 (2.2) |
| Prior MI (%) | 42 (2.1) | 19 (2.4) | 23 (1.9) |
| Heart failure (%) | 18 (0.9) | 12 (1.5) | 6 (0.5) |
| PVD (%) | 27 (1.3) | 13 (1.6) | 14 (1.2) |
| Stroke (%) | 1 (0.0) | 0 (0.0) | 1 (0.1) |
| COPD (%) | 71 (3.5) | 55 (6.9) | 16 (1.3) |
| CKD (%) | 22 (1.1) | 15 (1.9) | 7 (0.6) |
| Cancer (%) | 316 (15.8) | 167 (21.0) | 149 (12.3) |
| High risk for cardiovascular disease (%) | 403 (20.1) | 199 (25.1) | 204 (16.9) |
| High risk for lung cancer (%) | 235 (11.7) | 122 (15.4) | 113 (9.4) |
| High risk for breast or ovarian cancer (%) | 286 (14.3) | 101 (12.7) | 185 (15.3) |

BMI, body mass index; CKD, chronic kidney disease; COPD, chronic obstructive pulmonary disease; MI, myocardial infarction; PVD, peripheral vascular disease.

### Frequency of Urgent or Emergent Results

Among the 2,002 participants, 39.7% had at least one urgent (orange flag) or emergent (red flag) finding returned during or after the baseline visit, and 9.0% had at least two results returned (**Table 2**). Individuals with urgent or emergent results were older and had a greater comorbidity burden compared with those who did not have such results. For example, individuals with urgent or emergent results more frequently reported a history of cancer (21.0% vs. 12.3%), CAD (6.5% vs. 2.2%), or type 2 diabetes (20.0% vs. 8.6%) compared with those who did not have such findings.

### Types of Urgent and Emergent Results Returned

**Table 3** outlines, by testing modality, the specific urgent and emergent results that were returned to participants. The most common results returned from cardiac CT testing included incidental findings of pulmonary nodules (n=159); lung parenchyma findings (n=55), such as emphysema or fibrosis; and aortic aneurysms (n=22). The most common results returned from chest X-rays included pulmonary nodules (n=42); bone findings (n=28); and lung parenchyma findings (n=14), including atelectasis, emphysema, fibrosis, opacity, sarcoid, or infection. Among the 2,002 resting and stress echocardiography tests performed, 68 urgent or emergent results from resting echocardiograms were returned, and 123 urgent or emergent results from stress echocardiograms were returned. These results included valvular abnormalities or findings suggestive of ischemia on stress test. Most of the abnormal results on resting electrocardiograms were for bradycardia (n=74). The eye imaging results included 246 urgent or emergent findings related to eye abnormalities, including epiretinal membranes with foveal distortion or retinal hemorrhages. The most common urgent or emergent results from the laboratory tests included high hemoglobin A1c (n=56), abnormal vitamin D (n=46), and abnormal thyroid function (n=24). Additionally, 72 urgent or emergent results were reported for abnormal vital signs, such as bradycardia or high blood pressure.

**Table 3.** Number of Urgent and Emergent Incidental Findings by Modality*.

| **Modality/Most common findings** | **Urgent finding** | **Emergent finding** | **Total** |
| --- | --- | --- | --- |
| **Chest computed tomography** |  |  |  |
| Abdominal finding | 1 | 0 | 1 |
| Aortic aneurysm | 22 | 0 | 22 |
| Aortic finding, other than aneurysm | 3 | 1 | 4 |
| Bone finding | 2 | 14 | 16 |
| Breast abnormality | 13 | 0 | 13 |
| Hiatal or diaphragmatic hernia | 12 | 0 | 12 |
| Liver finding | 11 | 0 | 11 |
| Lung fluid finding | 1 | 0 | 1 |
| Lung parenchyma finding | 51 | 4 | 55 |
| Pericardial effusion | 9 | 0 | 9 |
| Pulmonary mass | 16 | 0 | 16 |
| Pulmonary nodule | 157 | 2 | 159 |
| Pneumothorax | 0 | 1 | 1 |
| Computed tomography other | 5 | 0 | 5 |
| **Chest X-ray** |  |  |  |
| Aortic aneurysm | 1 | 0 | 1 |
| Bone finding | 5 | 23 | 28 |
| Breast abnormality | 1 | 0 | 1 |
| Hiatal or diaphragmatic hernia | 5 | 0 | 5 |
| Lung fluid finding | 1 | 0 | 1 |
| Lung parenchyma finding | 14 | 0 | 14 |
| Pulmonary mass | 9 | 0 | 9 |
| Pulmonary nodule | 42 | 0 | 42 |
| Pneumothorax | 0 | 1 | 1 |
| Chest X-ray other | 3 | 0 | 3 |
| **Resting echocardiogram (echo)** |  |  |  |
| Abnormal resting echo** | 48 | 7 | 55 |
| Aortic aneurysm | 9 | 0 | 9 |
| Aortic finding, other than aneurysm | 1 | 0 | 1 |
| Pericardial effusion | 3 | 0 | 3 |
| **Stress echo** |  |  |  |
| Abnormal stress echo^ | 89 | 34 | 123 |
| **Resting electrocardiogram (EKG)** |  |  |  |
| Abnormal EKG - bradycardia | 48 | 26 | 74 |
| Abnormal EKG - other | 1 | 3 | 4 |
| **Eye imaging** |  |  |  |
| Abnormal eye findings | 244 | 2 | 246 |
| **Laboratory findings** |  |  |  |
| Abnormal cholesterol | 1 | 0 | 1 |
| Abnormal glucose | 9 | 0 | 9 |
| Abnormal renal function | 4 | 0 | 4 |
| Abnormal thyroid function | 24 | 0 | 24 |
| Abnormal vitamin D | 46 | 0 | 46 |
| High A1c | 56 | 0 | 56 |
| Abnormal hemoglobin | 0 | 1 | 1 |
| Abnormal white blood cells | 0 | 1 | 1 |
| **Vital sign findings** | **38** | **34** | **72** |
| Abnormal blood pressure | 23 | 10 | 33 |
| Bradycardia | 15 | 22 | 37 |
| Abnormal respiratory rate | 0 | 2 | 2 |
| **Total number of findings** | **1005** | **154** | **1159** |

\*Data presented in this table are on the level of the finding, rather than on the level of the patient.
\*\*Examples of abnormal resting echo: valvular abnormalities, structural abnormalities, functional abnormalities.
^Examples of abnormal stress echo: positive stress test (e.g. wall motion abnormalities, positive ECG findings).

Comparing all testing modalities, the most commonly returned results (urgent and emergent combined) were eye findings, pulmonary nodules, abnormal stress echocardiograms, abnormal rest electrocardiograms (bradycardia), and lung parenchyma findings, in that order. The demographics of the participants with those findings are presented in **Table 4**. Individuals with pulmonary nodule findings were more frequently current smokers (24.6% vs. 13.9%, p<0.001) and/or at high risk for lung cancer (19.8% vs. 10.9%, p=0.001) compared with those without pulmonary nodule findings. Participants with abnormal stress echocardiograms more frequently had a history of CAD (7.8% vs. 3.7%, p=0.05) or prior MI (5.2% vs. 1.9%, p=0.04) than those without abnormal findings. Individuals with lung parenchyma findings were more frequently Black or African American (29.7% vs. 17.5%, p=0.02), were current smokers (35.9% vs. 14.2%, p<0.001), had a history of COPD (25.0% vs. 32.8, p<0.001), and/or were at elevated risk for lung cancer (35.9% vs. 10.9, p<0.001) than those without lung parenchyma findings.

**Table 4.**
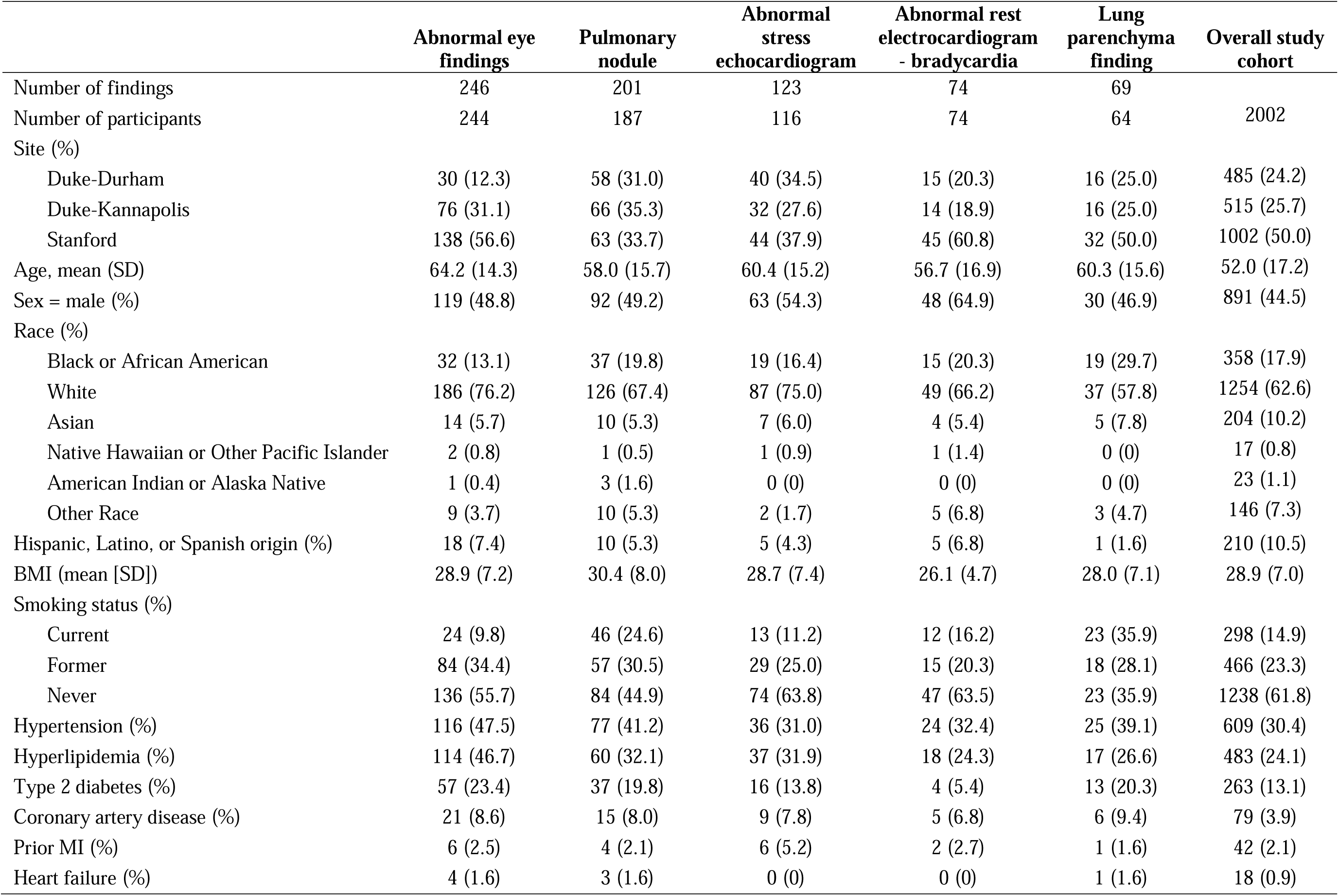
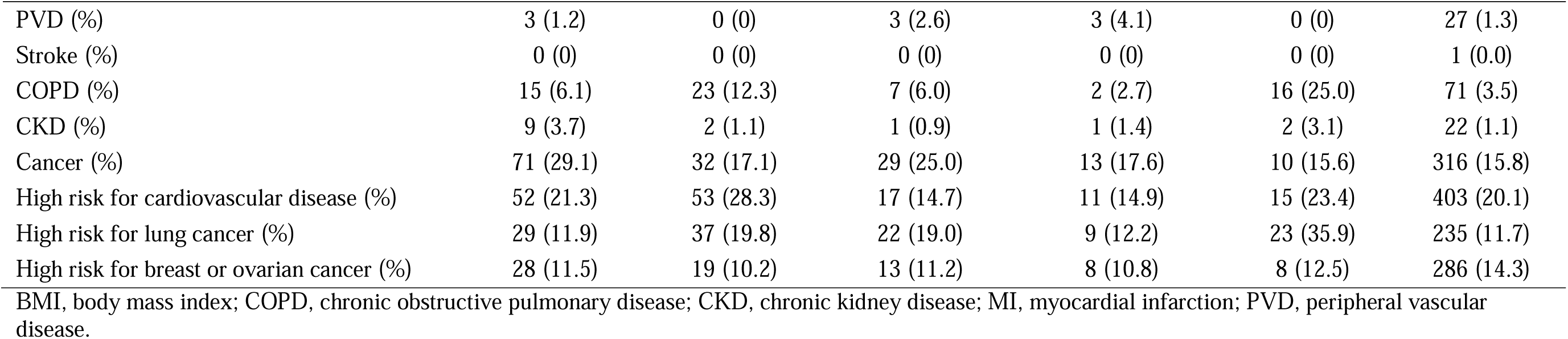
Demographic Characteristics and Medical History of Participants with Urgent or Emergent Findings.

### Model to Predict the Likelihood of Having Urgent or Emergent Results

The final model to discriminate between participants with vs. without urgent or emergent findings on the enrollment assessments included age; race; pulse; BMI; systolic blood pressure; total cholesterol; triglycerides; high-density lipoprotein (HDL) cholesterol; HbA1c; smoking status; enrollment site; and history of CAD, COPD, type 2 diabetes mellitus, MI, CKD, hypertension, or hyperlipidemia (**Supplementary Table 2**). The c-index of the final model was 0.70, with an accuracy of 67%. The calibration curve had an intercept of 0.86 and a slope of 1.06 in the test data compared with ideal values of 0 and 1, respectively. Similarly, the Hosmer-Lemeshow test *p*-value was 0.45, indicating no evidence for a lack of model fit.

## DISCUSSION

The PBHS study developed a comprehensive, systematic approach to returning individual-level urgent or emergent findings from multiple clinical modalities to research participants. We present herein: (1) our approach and experience with return of results; (2) a demonstration of the frequency of urgent or emergent incidental findings; and (3) the characteristics of participants with incidental findings, including a model to predict which individuals were most likely to have urgent or emergent findings. This roadmap has the potential to inform ongoing and future studies as we move collectively toward meeting the National Academies’ guidelines for return of results and democratization of data.

The results of this study demonstrate that urgent or emergent incidental findings, as defined in the PBHS study, are very common. In our study of 2,002 participants, 39.7% (n=794) had at least one urgent or emergent result, with abnormal eye findings and pulmonary nodules being the most common. Perhaps not surprisingly, individuals with urgent or emergent incidental findings tended to be older with a greater burden of cardiovascular, metabolic, or cancer comorbidities than those without urgent or emergent findings. Further follow-up of outcomes in the PBHS cohort will be necessary to understand the long-term health implications of these incidental research findings.

While prior and ongoing studies have taken a different approach, our experience with return of results highlights that urgent and emergent results, as defined by the PBHS study, require a significant effort from study clinicians and staff. The sheer number of urgent and emergent results returned to participants was much greater than anticipated. Furthermore, the process required considerable time from each of the clinicians who were returning results. Although some participants had more than one flagged finding, these results often became available at different times, necessitating separate contact points for each returned result. For example, some participants had up to 8 separate results returned. Future studies may want to consider more stringent criteria for urgent or emergent findings, which would lead to a smaller volume of results to return.

All emergent findings required direct contact with the participant, either in person at the study visit or over the phone, and the study clinician returning the result also generally helped to arrange or facilitate urgent follow-up for the participant. While urgent results could be returned via the EHR if participants had created their own EHR account, many were returned via phone call. In part, this was because many participants had questions about their results, and some had significant anxiety around their abnormal tests, which were better addressed over the phone. In future studies, ensuring that participants allow their personal clinicians to be contacted by study staff may help to alleviate both the anxiety of participants and the burden on study staff. However, this would potentially increase the burden on the participants’ clinicians, which would need to be closely evaluated and weighed.

The PBHS effort to return individual-level results reflects a movement toward viewing participants as partners in research rather than subjects. Indeed, many studies have clearly shown that research participants want their results to be returned to them: a review of 8 studies that assessed participants’ attitudes toward receiving individual-level results revealed that the overwhelming majority wished to receive such results (7). Importantly, this finding was consistent across types of clinical research and disease states. The desire to receive individual-level results may reflect one of the expectations that people have when participating in research, which is to improve one’s own health (8). Participants may also desire to have information that is relevant for family members or they may believe that the return of individual-level results is the right of participants and shows respect (7). Therefore, it is important that plans and investment in resources for return of results are made at the study design phase.

Despite increasing calls for return of results, minimal guidance has been published on how to accomplish this process. In genetics, multisite research programs, including the Clinical Sequencing Exploratory Research (CSER) Consortium and the Electronic Medical Records and Genomics (eMERGE) Network, have developed practical strategies to return genetic data to participants (9). Other groups have also published recommendations and consensus statements to guide the return of genetic data (10-12). Additionally, environmental health biomonitoring science has provided optimal methods to communicate results to participants (13,14). However, outside of these fields, recommendations for return of results are sparse. In response to a request by the National Institutes of Health (NIH), Centers for Medicare and Medicaid Services, and Food and Drug Administration, the National Academies released guidance on returning individual-level research results to participants (5). While this report clearly endorses return of results in principle, minimal details are given on exactly how to carry this out (i.e. which results, when, how, and by whom should they be returned). Even less is known about the impact of return of results on study investigators and study resources.

Several caveats should be considered when interpreting our work. This is the experience of a single study, and as such, the frequency and types of flagged results may not be generalizable to other studies or populations. The PBHS study performed extensive imaging, including eye imaging, which was beyond the testing typically conducted in studies. These tests resulted in a larger burden of urgent and emergent findings. While urgent and emergent results were partially defined by clinical guidelines, some determinations could not be made based on the guidelines given the extensive phenotyping and lack of guidelines in this space. Therefore, our RoR Committee protocols could serve as a basis for future studies to consider in their return of results protocols. The exact time spent by each clinician returning results was not recorded, nor was clinician perception of the process. Additionally, we do not yet know how participants perceived the return of results process, what they did with the information, and how it impacted their health care; these studies are currently underway.

The scientific community’s understanding of the return of individual-level, clinical (i.e. non-genetic or non-environmental) results to research participants is still in its infancy. Indeed, much is still unknown about the overall utility of returning results to participants, the effects on various stakeholders, and optimal ways to conduct the process. Specifically, whether and how participants utilize the returned information and the impact on participant engagement in research are important gaps in knowledge. Furthermore, the impact of returning results can vary by age, gender, race and ethnicity, and socioeconomic status. A critical question is whether returning results can increase engagement in clinical research from traditionally under-represented groups. In addition, the process of returning results must be streamlined and optimized. More research is required to understand whether individual clinical results can or should be returned by study personnel other than clinicians, whether it can be done entirely through digital means, and whether certain modalities are preferred by participants over others. Because participants may prefer different methods, efforts should be made to understand how best to tailor the process of returning results by study, by result type, and most importantly, by participant.

Given these critical unanswered questions, we advocate for the collaborative development of a research agenda that is focused on optimizing the return of individual-level clinical results in research studies. Such an endeavor will require recognition and support from funding agencies, such as the NIH and Patient-Centered Outcomes Research Institute. Stakeholders, including researchers, participants, and funders, will need to co-develop this research agenda to ensure that all viewpoints are represented. Only then can the return of results process elevate clinical research as a true partnership between participants, funders, and researchers.

## Conclusions

The PBHS study is one of the first clinical research studies to systematically and rigorously return a wide variety of individual-level clinical results to research participants. The process outlined herein led to the return of clinically important test results but was resource-intensive. The return of results is an important component of the synergistic relationship between researchers and participants and is the subject of increasing attention. However, much remains unknown about how the return of results impacts both participants and research programs and how this process can be streamlined and optimized according to participant preferences. This will require a concerted effort to develop a research agenda around the return of results that includes the perspectives of participants, researchers, and funders alike. In the meantime, the PBHS experience can serve as a guide for researchers committed to returning individual-level clinical results to participants.

## Data Availability

The deidentified Project Baseline Health Study (PBHS) data corresponding to this study are available upon request for the purpose of examining its reproducibility. Requests are subject to approval by PBHS governance.

## Acknowledgments

The authors wish to thank Project Baseline Health Study participants and study sites.

## Ethics Statement

The study was funded by Verily Life Sciences and was coordinated by the Duke Clinical Research Institute. The study was approved by the Duke University and Stanford University Institutional Review Boards. Participants provided written informed consent before participation, and the study observed the privacy rights of all participants.

## Author Contributions

*Conceptualization:* RCG, SS, NP

*Formal Analysis:* GJJ, RCG, SG, LK, SS, NP

*Methodology:* SG, LK, SS, NP

*Project Administration:* BH, JE, EF

*Supervision:* AH, KM, SS, RCG

*Writing − original draft:* NP, SS

*Writing − review & editing:* All authors.

All authors had access to the data and contributed to the preparation of this manuscript.

## Abbreviations

CHLI: California Health and Longevity Institute
CKD: chronic kidney disease
CLIA: Clinical Laboratory Improvement Amendments
COPD: chronic obstructive pulmonary disease
CSER: Clinical Sequencing Exploratory Research
CT: computed tomography
EHR: electronic health record
MI: myocardial infarction
PBHS: Project Baseline Health Study
PI: principal investigator
PVD: peripheral vascular disease
RoR: return of results

