## Supplemental Material for "Returning Individual-Level Urgent or Emergent Research Results to Participants: The Project Baseline Health Study Experience"

**Supplemental Table 1. Clinical Laboratory Emergent and Urgent Findings**

|  | **Urgent findings** | **Emergent findings** |
| --- | --- | --- |
| **Hematology** |  |  |
| Hematocrit (%) | n/a | <20.1 OR >59.9 |
| Hemoglobin (g/dL) | n/a | <7.1 OR >19.9 |
| Platelets (thousand) | n/a | <31 OR >1,499 |
| White blood cells (thousand/mcl) |  | <1.1 OR >50 |
| Manual differential | Blasts of malignant cells | n/a |
| **Chemistry** |  |  |
| HbA1c (%) | ≥9 | n/a |
| Calcium (mg/dL) | n/a | <6 OR >13 |
| Glucose (mg/dL) | 300-500 | <40 OR >500 |
| Potassium (mEq/L) | n/a | <2.8 OR >6 |
| Sodium (mEq/L) | n/a | <120 OR >160 |
| AST (U/L) | ≥500 | n/a |
| ALT (U/L) | ≥500 | n/a |
| Magnesium (mEq/L) | n/a | <0.7 OR >5 |
| Serum creatinine (mg/dL) | >2 | n/a |
| LDL-c (mg/dL) | >220 | n/a |
| Triglycerides (mg/dL) | >1000 | n/a |
| TSH (mcIU/ML) | >50 OR <0.1 | n/a |
| 25-OH Vitamin D (ng/mL) | <10 | n/a |
| Phosphorus (mg/dL) | n/a | <1.0 |
| Amylase (U/L) | >500 | n/a |
| Lipase (U/L) | >200 | n/a |
| Carbon dioxide (mEq/L) | n/a | <15 |

ALT, alanine transaminase; AST, aspartate transaminase; HbA1c, glycated hemoglobin; LDL-c, low-density lipoprotein cholesterol; OR, odds ratio; TSH, thyroid-stimulating hormone.

**Supplemental Table 2. Significant Predictors of Red or Orange Incidental Findings in the LASSO Model**

| **Variable** | **Coefficient in LASSO model** |
| --- | --- |
| Age | 0.021 |
| White race | 0.029 |
| Heart rate | -0.013 |
| BMI | 0.00086 |
| Systolic BP | 0.0067 |
| Total cholesterol | -0.00012 |
| Triglycerides | 0.00017 |
| HDL | -0.000043 |
| HbA1c | 0.15 |
| Smoking status: non-smoker | -0.11 |
| History of CKD | 0.85 |
| History of hypertension | 0.14 |
| History of hyperlipidemia | 0.11 |
| History of CAD | 0.016 |
| History of COPD | 0.80 |
| History of DM2 | 0.52 |
| History of MI | -0.21 |
| Enrollment site: Duke-Kannapolis | 0.039 |

BMI, body mass index; BP, blood pressure; CAD, coronary artery disease; CKD, chronic kidney disease; COPD, chronic obstructive pulmonary disease; DM2, type 2 diabetes mellitus; HDL, high-density lipoprotein; HbA1c, glycated hemoglobin; MI, myocardial infarction.
